# Prognostic value of tumor-informed ctDNA in HPV-independent head and neck squamous cell carcinoma

**DOI:** 10.64898/2026.01.09.26343802

**Authors:** Daniel A. Ruiz-Torres, Thomas J. Roberts, Pan Du, Julia Mendel, Saskia Neagele, Giancarlo Bonora, Frank Zhang, Vasileios Efthymiou, Ross D. Merkin, Derrick T. Lin, Jonathan J. Paly, Mark A. Varvares, Daniel G. Deschler, Allen L. Feng, Jeremy D. Richmon, Adam S. Fisch, Shidong Jia, Daniel L. Faden

## Abstract

**Importance:** Recurrence rates for locally advanced HPV-independent (HPV-) head and neck squamous cell carcinoma (HNSCC) are high. Circulating tumor DNA (ctDNA)-based minimal residual disease (MRD) assays have shown promise to improve management and surveillance in several tumor types, but their clinical utility in HPV- HNSCC remains understudied.

**Objective**: To evaluate the performance of a tumor-informed ctDNA-based MRD assay (PredicineBEACON) in patients with newly diagnosed, locally advanced HNSCC (LA-HNSCC).

**Design:** Between 12/2020 and 3/2022 ctDNA was assessed before surgery, before the start of adjuvant treatment (MRD-E), within six weeks of completion of treatment (MRD-TC), and during surveillance (MRD-S). Patients were followed for at least 12 months after treatment completion. We used Kaplan-Meier survival analyses to compare recurrence-free survival (RFS) and overall survival (OS) between patients who were MRD positive those who were MRD negative during each time window. Multivariable Cox hazard regressions were used to assess the association between MRD status and outcomes while controlling for established risk factors.

**Setting**: This was a prospective cohort study of patients treated at a large referral center specializing in treatment of HNSCC.

**Participants**: Forty patients with newly diagnosed, LA-HNSCC treated with surgery followed by risk-adjusted adjuvant treatment

**Intervention**: Tumor-informed ctDNA-based MRD testing

**Main Outcomes and Measures**: Recurrence-free survival (RFS) and overall survival (OS)

**Results:** We processed 142 samples from 40 patients. The median age was 63, 27% were female, 87% were Caucasian, and 95% had HPV- disease. Fifty percent (20/40) of patients experienced recurrence. The pre-surgery ctDNA detection rate was 97% (35/36). MRD-TC positivity was associated with worse OS (HR= 7.15; 95% CI: 1.44-35.3) and RFS (HR= 5.39; 95% CI: 1.98-21.07). MRD-S positivity was associated with worse RFS (HR=8.2; 95% CI: 2.06-33.6). The median time from first MRD detection to clinical detection of recurrence was 5 months (range 0.2-21.6). In multivariable analyses, MRD positivity was associated with worse RFS (HR 13.8; 95% CI 2.92-65.7) and worse OS (HR 18.9; 95% CI 2.27-158).

**Conclusions and Relevance:** Tumor-informed ctDNA MRD positivity was associated with worse RFS and OS in patients with HNSCC. MRD testing could serve as a non-invasive, prognostic biomarker in HPV- HNSCC patients.

**Key Points:** **Question**: Among patients with locally advanced HNSCC (LA-HNSCC) treated with curative-intent surgery, is minimal residual disease (MRD) detection using a tumor-informed ctDNA-based assay prognostic for recurrence free survival (RFS) or overall survival (OS)?

**Findings**: In this prospective cohort study of 40 patients with LA-HNSCC, nearly all of whom had HPV-independent disease, MRD positivity during the first 6 weeks after completing treatment and during surveillance was associated with worse OS and RFS.

**Meaning**: Tumor-informed ctDNA MRD detection after treatment completion could serve as a non-invasive, prognostic biomarker in HPV-independent HNSCC patients.

**Social Media Post:** Findings by @FadenLab and colleagues show that detection of MRD using a tumor-informed ctDNA-based after treatment completion is associated with worse recurrence free survival and overall survival in HPV-independent HNSCC.

## Introduction

Head and neck squamous cell carcinomas (HNSCC), a term to describe tumors of the oral cavity, oropharynx, hypopharynx, larynx, nasopharynx, and sinonasal spaces, are collectively the seventh most common cancer worldwide^1^. More than 60% of cases of HNSCC are diagnosed as locoregionally advanced disease and are often treated with surgery followed by radiation or concurrent chemoradiation (CRT) based on the risk of recurrence as assessed by surgical pathology^2,3^. While human papilloma virus (HPV)-associated (HPV+) tumors have high cure rates, recurrence rates remain high among patients with HPV-independent (HPV-) HNSCC. Approximately 26-50% of patients with stage III and non-metastatic stage IV HPV- HNSCC will develop recurrence^4,5^. Current approaches to post-treatment surveillance rely on frequent invasive physical exams and imaging, and treatment options for recurrent HNSCC are limited—patients often require morbid salvage surgeries or treatment with systemic therapies that have substantial toxicity and response rates below 50%^6–9^. Non-invasive prognostic biomarkers for patients with HNSCC have the potential to inform adaptive treatment intensification and deintensification approaches, as well as to help detect recurrent disease earlier during post-treatment surveillance.

Circulating tumor DNA (ctDNA) is shed into the bloodstream by tumor cells in patients with cancer^10^. The detection of ctDNA following curative-intent therapy, commonly referred to as molecular residual disease (MRD), is strongly associated with patient outcomes across solid cancer types^11–14^. There are two main types of MRD assays: non-personalized (tumor-naïve), and personalized (tumor-informed). The latter requires tumor tissue but provides higher sensitivity. Emerging strategies are integrating methylome profiling in tumor-naive assays and employing multiparametric tests powered by AI-based algorithms, obtaining promising results^15^. CtDNA approaches to detect MRD have demonstrated prognostic value across multiple tumor types and can detect recurrence earlier than conventional surveillance methods^11^. Among patients with HPV+ HNSCC, ctDNA-based assays have shown improved post treatment surveillance, diagnostic accuracy, lower costs, and shortened time to diagnosis of recurrence^16,17^. However, data on HPV- HNSCC remains limited. Given the high incidence of recurrent disease and its associated morbidity, more data on the performance of MRD assays in patients with HPV-HNSCC are needed.

Here, we present a prospective cohort study of patients with HPV- HNSCC to evaluate the performance and prognostic value of PredicineBEACON, a validated tumor-informed MRD detection assay with published limits of detection of 0.0025% variant allele frequency (VAF). Our primary hypothesis was that MRD detection following treatment completion would be associated with elevated risk of recurrence and poorer survival outcomes.

## Methods

### Study population

Patients were eligible for this study if they were diagnosed with locally advanced non-nasopharynx HNSCC (oral cavity, oropharyngeal [OPC], larynx, or hypopharynx) and underwent surgery at Massachusetts Eye and Ear between December 2020 and July 2023. The determination of locally advanced disease was made by the treatment team independent of stage. Selected patients either had pathologically confirmed HPV- HNSCC or were felt prior to surgery to have a high probability of having HPV- HNSCC. Formalin-fixed paraffin-embedded (FFPE) tumor tissue was required for enrollment, with a head and neck pathologist (ASF) reviewing all tumor tissue to ensure sufficient tumor quantity and quality for molecular testing. Patient characteristics were captured via chart review, and patient race was based on self-reported data in the EHR. This study was approved by the Dana-Farber/Harvard Cancer Center Institutional Review Board (DFCI 18-653), and all patients provided written informed consent.

### Clinical specimens

Plasma samples were collected before surgery when logistically feasible. Among patients who were treated with surgery alone (referred as ‘SURG’ across the manuscript), subsequent plasma samples were collected at surgical follow-up appointments (Figure 1A). Among patients who received either post-operative radiation or CRT, hereinafter referred to as ‘adjuvant patients,’ plasma specimens were collected during clinic visits before and after post-operative treatment and during post-treatment surveillance. All plasma specimens were collected at visits scheduled as part of patients’ routine clinical care. No additional visits were scheduled specifically for plasma collection, which resulted in some patients not having samples drawn during some windows. Blood samples were drawn in EDTA tubes, and plasma isolation was obtained after two rounds of centrifugation (10 mins at 1600 RCF and 3000 RCF). Plasma and buffy coat were then aliquoted and stored at –80°C until processing.

**Figure 1.**
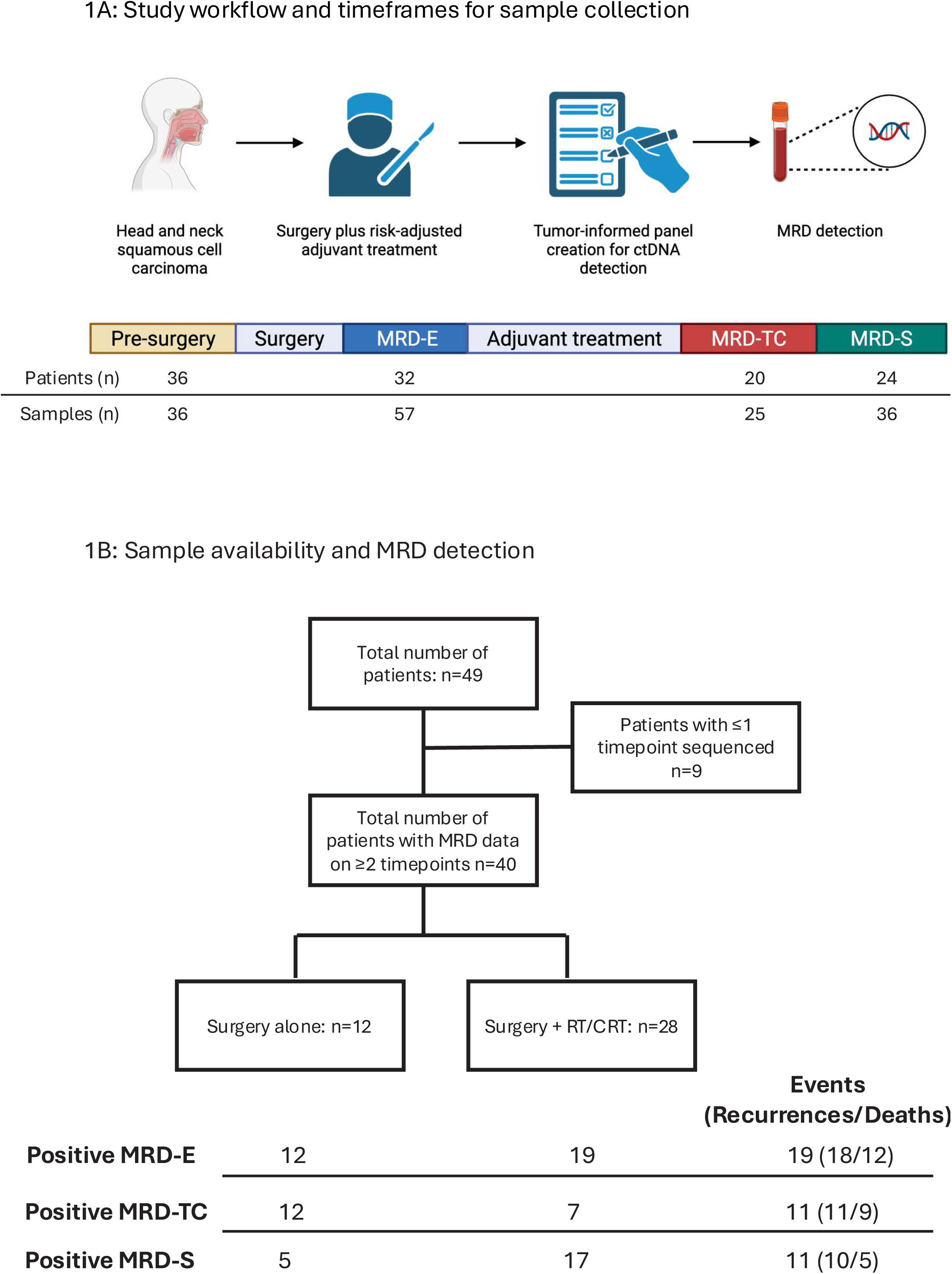
Study workflow. (A) Flow diagram illustrating the overall study design and sample collection timeline: pre-surgery, then from postoperative day 1 to the start of adjuvant therapy (MRD-E), up to six weeks after completion of adjuvant therapy (MRD-TC), and from more than six weeks to six months post-adjuvant therapy (MRD-S). (B) The flow chart shows the total number of patients included in the study, followed by the treatment received, recurrence, and MRD status.

### MRD Testing

Tumor tissue and plasma samples were processed at Predicine Laboratories utilizing PredicineBEACON, a previously reported NGS-based tumor-informed assay^18–20^. This assay utilizes PredicineWES+, a boosted whole-exome sequencing (WES) on tumor specimens to evaluate 20,000 genes across the whole exome in addition to the boosted sequencing of 600 cancer-related genes for baseline variant profiling^13,19,21,22^. For each patient, a tumor-specific MRD panel is designed to include up to 50 personalized somatic mutations along with a fixed panel covering 500 actionable and hotspot mutations. After the creation of the tumor-specific panel, the assay applies ultra-deep sequencing (100,000x) to plasma samples to identify genomic alterations included in the MRD panel. In addition, the PredicineSCORE copy number burden (CNB) assay with a ∼3X low-pass whole genome sequencing (LP-WGS) was performed for both baseline and on-treatment plasma samples. The MRD calling was reported as a binary outcome (positive/negative) based on previously used criteria using mutation and copy number status. A specimen was classified as positive if two or more monitored alterations were detected with an estimated tumor fraction higher than 0.00001. The assays were performed in a CLIA-certified and College of American Pathologists–accredited laboratory (Predicine Inc., Hayward, CA).

### Clinical data

Clinical data were censored on 6/13/2024 and obtained from chart reviews completed between 7/1/2024 and 12/31/24 to obtain pathology details, treatments, treatment dates, and the outcomes of interest, recurrence-free survival (RFS) and overall survival (OS). Charts were reviewed independently by DRT and TJR, and differences were resolved by consensus. Investigators were blinded to the MRD results when performing chart reviews. When reporting alteration frequency, we reported genomic alterations that were classified as “pathogenic” or “likely pathogenic” in the National Center for Biotechnology Information’s ClinVar database.

### Statistical analysis

This study evaluated MRD detection during three-time windows: early after surgery (MRD-E), after treatment completion (MRD-TC), and during surveillance (MRD-S). The window for MRD-E was defined as the first 6 weeks after surgery for surgery-only patients and the period between surgery and completion of radiation or CRT for patients who received adjuvant therapy. The MRD-TC window encompassed the first 6 weeks after surgery for surgery-only patients or the first 6 weeks after completion of adjuvant therapy for patients who received adjuvant therapy. The MRD-S window included all time points after the MRD-E and MRD-TC periods.

For each of these windows, we calculated the sensitivity and specificity at the patient level and at the test level to evaluate test performance. When calculating at the patient level, MRD-positive was defined as a patient having any MRD test that was positive during the specified window (e.g. a patient with a positive and a negative test during a window was defined as positive for that window). Patients were classified as MRD-negative during a given window only if all results during the window were negative. A true negative value was defined as no evidence of clinically identifiable disease prior to the date of data censoring. Kaplan-Meier survival analyses were used to evaluate the association between MRD detection during the specified windows and OS and RFS.

Univariable and multivariable Cox proportional hazard models were used to assess the association between MRD detection and OS and RFS. To avoid overfitting models, the data from the MRD-TC and MRD-S windows were pooled for multivariate models for OS and RFS. This approach increased the number of patients and events included in the models. In these models, any positive MRD result during the MRD-TC or MRD-S window was coded as MRD positive. Due to the low number of events, covariates included in these models were limited to two binomial variables: presence of > 1 mm of extranodal extension (ENE), and margin status (positive or negative). Patients were classified as having involved margins if the pathology report indicated tumor was present at or within 2 mm of the specimen margin. Values were considered statistically significant if α < 0.05. All analyses were performed using R Statistical Software (v4.3.1, R Core Team 2023). The code used for the analysis and plotting is available upon reasonable request from the corresponding author.

## Results

### Study population & sample collection

This study included 40 patients who had at least two samples processed during the period of analysis (Figure 1B). The median follow-up was 33.7 months (range 7.12-40.4). Baseline characteristics are summarized in Table 1. Patients were mostly White (87%), and male (73%). The median age at diagnosis was 63 (range 28-85), and most had a history of tobacco exposure (55%). Nearly all patients (95%) had HPV- disease. Most malignancies were oral cavity tumors (65%), 87% had T3 or T4 disease, and 43% had pathologic nodal involvement. Seventy percent of patients received post-operative radiation (20%) or CRT (50%). When looking at high-risk features on pathology, 17% of patients had ENE, and 50% of patients had involved margins.

**Table 1.** Clinical characteristics for the entire cohort

| Parameter | All (n=40) (%) | MRD-E (+)<br>(n=17/31) | MRD-TC (+)<br>(n=9/19) | MRD-S (+)<br>(n=8/22) |
| --- | --- | --- | --- | --- |
| Age, y (median, range) | 63 (28-85) | 66 (48-81) | 65 (48-78) | 66 (49-83) |
| **Sex** |  |  |  |  |
| Female | 11 (27) | 3 | 3 | 0 |
| Male | 29 (73) | 14 | 6 | 8 |
| **Race** |  |  |  |  |
| Asian | 2 (5) | 1 | 2 | 0 |
| Black | 0 | 0 | 0 | 0 |
| White | 35 (87) | 16 | 7 | 8 |
| Declined | 2 (5) | 0 | 0 | 0 |
| Not reported | 1 (2.5) | 0 | 0 | 0 |
| **Ethnicity** |  |  |  |  |
| Not Hispanic | 37 (92) | 16 | 8 | 7 |
| Hispanic | 1 (2) | 0 | 0 | 0 |
| Declined | 1 (2) | 0 | 0 | 0 |
| Not reported | 1 (2.5) | 1 | 1 | 1 |
| **Smoking status** |  |  |  |  |
| Former | 22 (55) | 9 | 5 | 4 |
| Never | 15 (37) | 6 | 4 | 4 |
| Current | 3 (7) | 2 | 0 | 0 |
| **Primary disease site** |  |  |  |  |
| Oral cavity | 26 (65) | 10 | 6 | 6 |
| Oropharynx | 5 (12) | 2 | 2 | 2 |
| Larynx | 6 (15) | 5 | 1 | 0 |
| Hypopharynx | 3 (7) | 2 | 1 | 0 |
| **HPV status determination** |  |  |  |  |
| p16 and/or HPV RNA ISH positive | 2 (5) | 2 | 1 | 0 |
| p16, HPV RNA ISH and/or PCR negative | 38 (95) | 15 | 8 | 8 |
| **Pathological T staging** |  |  |  |  |
| T0 | 0 | 0 | 0 | 0 |
| T1 | 1 (2) | 0 | 0 | 0 |
| T2 | 4 (10) | 1 | 0 | 3 |
| T3 | 16 (40) | 7 | 4 | 2 |
| T4 | 19 (47) | 9 | 5 | 3 |
| **Pathological N staging** |  |  |  |  |
| N0 | 23 (57) | 9 | 5 | 4 |
| N1 | 1 (2) | 0 | 0 | 0 |
| N2 | 6 (15) | 2 | 1 | 2 |
| N3 | 7 (17) | 4 | 2 | 1 |
| Unable to assess | 3 (7) | 2 | 1 | 1 |
| **Metastatic disease** |  |  |  |  |
| M0 | 40(100) | 17 | 9 | 8 |
| M1 | 0 | 0 | 0 | 0 |
| **Overall staging** |  |  |  |  |
| I or II | 5 (12) | 1 | 1 | 2 |
| III, IV A or IVB | 35 (87) | 16 | 8 | 8 |
| IVC | 0 | 0 | 0 | 0 |
| **Disease status at the time of the initial testing** |  |  |  |  |
| Incident | 26 (65) | 11 | 4 | 4 |
| Recurrent | 14 (35) | 6 | 5 | 4 |
| **Treatment modality** |  |  |  |  |
| Surgery only | 12 (30) | 7 | 7 | 2 |
| POP RT | 8 (20) | 3 | 0 | 1 |
| POP CRT | 20 (50) | 7 | 2 | 5 |
| **Margins** |  |  |  |  |
| Margins (+) | 20 (50) | 10 | 8 | 2 |
| Margins (-) | 18 (44) | 5 | 1 | 6 |
| Margins NA | 2 (5) | 2 | 0 | 0 |
| **ENE** |  |  |  |  |
| ENE (+) | 7 (17) | 3 | 1 | 2 |
| ENE (-) | 30 (75) | 12 | 7 | 5 |
| ENE NA | 3 (7) | 2 | 1 | 1 |

HPV = Human Papilloma Virus, RNA = ribonucleic acid, ISH = in situ hybridization, PCR = polymerase chain reaction, POP: Post-operative; RT: radiation therapy. CRT: Chemoradiation. ENE: Extra-nodal extension.

One hundred forty-two samples were tested, 3.6 samples per patient (range: 2–6) (Figure 2C). Specimens were collected for 36 patients pre-surgery. During the MRD-E window, 57 specimens were collected among 32 patients. During the MRD-TC window, 25 specimens were collected among 20 patients. During the MRD-S window, 36 specimens were collected among 24 patients (Figure 1A). For most instances where a patient did not have a specimen collected during a specific window, including the pre-surgery window, the reason a sample was not collected was due to the logistics of samples only being collected at routine clinic visits at one clinical site.

**Figure 2.**
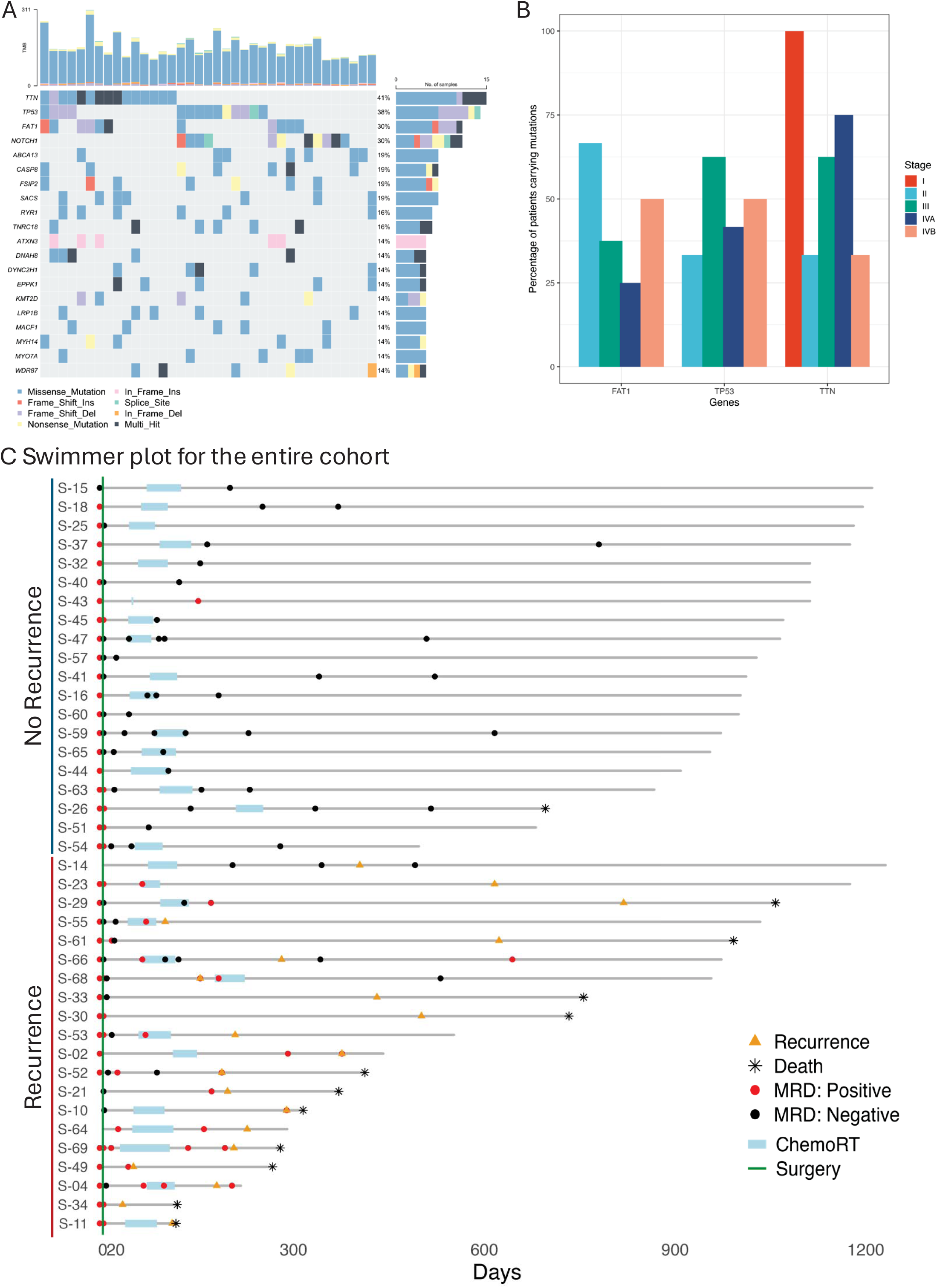
Genomic landscape and longitudinal MRD dynamics. (A) Oncoplot displays the top 20 pathogenic genomic alterations in the tissue samples for the cohort. (B) Bar plot showing the percentage of patients’ tumors harboring pathogenic alterations in genes *FAT1*, *TTN*, and *TP53* based on stage. (C) Swimmer plot illustrating the longitudinal dynamics of MRD detection for the entire cohort.

### Analysis of biomarkers in surgical tissue samples

The assay identified 2459 somatic mutations and 208 gene copy number variations across all patients. The most common alterations that were classified as pathogenic are shown in Figure 2A. The median tumor mutation burden score was 1.3 (range 0.1- 8.1). The most frequently altered genes included *TTN* (41%), *TP53* (38%), *FAT1* (30%), and *NOTCH1* (30%), as expected^23^. Gene copy-number losses were mostly observed in *TP53.* Among patients with *TP53* variations, three included *TP53* gene losses, whereas 22 had mutations. No meaningful difference was found between the prevalence of *TP53* variations based on stage (Figure 2B). No meaningful difference in survival outcomes was observed based on the presence of TP53 mutations [HR = 0.6; 95%CI: 0.2 - 2.24] (Supp. Fig. 1A).

### MRD detection rate and test performance

The pre-operative ctDNA detection rate was 97% (35/36). The median tumor fraction (TF) was 0.00822; range: 0.00016-0.08084). The baseline TF was evenly distributed across tumor locations, T, N, or AJCC stages (Supplemental Figure 1B-E). However, there was a moderately strong positive correlation between AJCC 8t stage and pretreatment TF (Pearson R= 0.4) (Supplemental Figure 1F). This association remained positive after adjusting for age, and smoking. Among patients with a tumor fraction higher than the median, there was a greater reduction in overall survival (OS) compared to patients with a tumor fraction lower than the median [HR: 1.9; 95% CI: 0.52 to 6.83]. However, the wide confidence interval indicates considerable imprecision, preventing definitive conclusions (Supplemental Figure 1G).

During the MRD-E window, samples were obtained at a median of 1 day (range: 1-64) after surgery for patients who received adjuvant CRT and at a median of 1 day (range: 1-40) for the patients who received surgery only. During the MRD-E window, 56% (18/32) of patients had a positive MRD results. Rates of detection were 60% (12/20) among patients who received adjuvant therapy and 58% (7/12) among patients treated with surgery alone. Of the 18 patients who were MRD-E positive, 13 (72%) experienced recurrences (ADJ: 7; SURG: 6), leading to a sensitivity of 77%, specificity of 67% (positive predictive value (PPV): 72%; negative predictive value (NPV): 71%). When calculating test characteristics at the test level, the MRD-E positivity had a sensitivity of 55%, specificity of 75% (PPV: 70%; NPV: 62%) for recurrence (Supplemental Table 1).

During the MRD-TC window, samples were obtained at a median of 6 days (range: 1-40) after completion of treatment (SURG: median of 1 day (range: 1-40); ADJ: median of 6 days (range 5-35)). During the MRD-TC, 40% (8/20) of patients had a positive MRD result. (ADJ 2/9, 22%; SURG: 6/11, 55%). Of the 8 patients who had an MRD-positive result during the MRD-TC window, 7 (88%) recurred (ADJ: 2; SURG: 5). This resulted in a sensitivity of 78%, specificity of 91% (PPV: 88%; NPV: 83%) (Supplemental Table 1). At the test level, MRD-TC detection had a sensitivity of 64%, specificity of 93% (PPV: 88%; NPV: 77%). The one false positive during the MRD-TC window was on post-op day (POD) 1 in a surgery only patient, suggesting POD 1 pay be too soon to assess for MRD with a highly sensitive assay. The lead time of MRD-TC detection to clinical detection of recurrence was at a median of 157 days (range: 8-649).

During the MRD-S window, samples were obtained at a median of 78 days (range: 6-186) after completion of treatment (SURG: median of 102, range 72-171; ADJ: median of 78, range: 6- 186). During the MRD-S window 42% (10/24) of patients had a positive MRD test (ADJ: 7/19, 37%; SURG: 3/5, 60%). Out of the 10 patients with positive MRD-S detection, 8 experienced recurrences (ADJ 6, SURG: 2). This resulted in a sensitivity of 89%, specificity of 87% (PPV: 80%; NPV: 93%). In contrast, at a test level, MRD-S detection led to a sensitivity of 67%, specificity of 90% (PPV: 83%; NPV: 78%) (Supplemental Table 1). The lead time of MRD-S detection to clinical detection of recurrence was at a median of 46 days (range:0-85) (Figure 2C). Notably, for patient S-04, the sample available during the MRD-S period was 24 days after clinical detection of recurrence. However, the sample prior to that one obtained at the end of adjuvant treatment was positive.

### MRD detection and survival

At a median follow-up of 33.7 months (range 12-40) (Supp. Fig 2B), the median OS for the entire cohort was not reached. One-year OS was 87% (95% CI 76.8-97.6). The median RFS for the cohort was 27 months (range:1.0 –39.7), and one-year RFS was 67% (95% CI 53-82). RFS and OS were compared for patients with detectable versus undetectable MRD for each timeframe. Among patients with positive MRD-E, there was a greater reduction in OS compared to patients with negative MRD-E [HR 2.86;95% CI 0.84 - 9.7]. However, the wide confidence interval indicates considerable imprecision, preventing definitive conclusions (Supplemental Figure 2A). In univariable analysis, OS and RFS were worse for MRD-TC positive patients compared to MRD-TC negative patients (OS: HR 7.15, 95% 1.44-35.3; PFS: HR 5.39, 95% 1.98-21.07) (Figure 3A). Similarly, patients with positive MRD-S had lower OS (HR 9.18, 95% CI 0.85-32.4) and worse RFS (HR 8.2, 95% 2.03-33.6) (Figure 3B).

**Figure 3.**
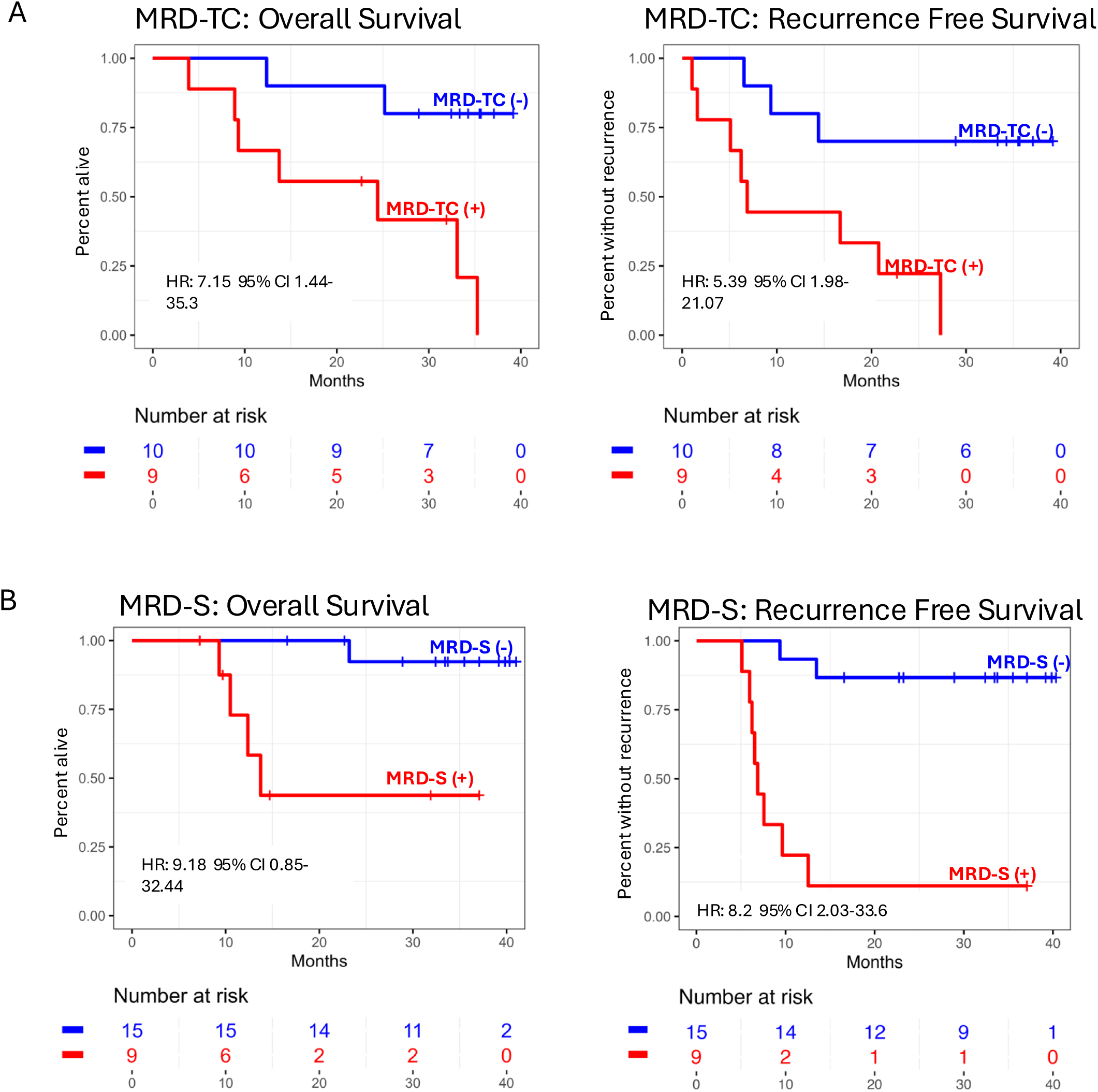
MRD detection impacts overall and recurrence-free survival. Detection of MRD-TC (A) and MRD-S (B) is associated with worse 3-year overall and recurrence-free survival in univariable analysis.

In multivariable analyses, detection of MRD was associated with worse RFS (HR: 13.1; 95% CI 2.92-65.7) and OS (HR 18.9; 95% CI 2.27-158) after adjusting for ENE and margin status (Table 2). Supplementary Figure 2B shows overall survival and recurrence curves for entire cohort and Figures 2C-E shows survival and recurrence as a function of ENE, margin status and AJCC stage.

**Table 2.** Multivariable Cox proportional hazard regression analyses.

| Variable | Overall Survival |  | Recurrence Free Survival |  |
| --- | --- | --- | --- | --- |
|  | aHR | 95% CI | aHR | 95% CI |
| MRD | 18.9 | 2.27-158 | 13.8 | 2.92-65.7 |
| ENE | 0.94 | 0.18-4.97 | 0.84 | 0.22-3.16 |
| Positive margins | 1.3 | 0.31-5.39 | 0.82 | 0.26-2.63 |

aHR = adjusted Hazard Ratio, CI = confidence interval, MRD = detectable minimal residual disease during treatment completion or surveillance windows, ENE = extranodal extension

## Discussion

This study evaluated the performance of a tumor-informed whole-exome-based MRD assay at multiple time points in a cohort of patients with mostly HPV- HNSCC who received curative intent surgery followed by risk-adjusted adjuvant therapy. We demonstrated that the detection of MRD using a ctDNA-based assay early after treatment was associated with worse OS and RFS, even after controlling for traditional risk factors for recurrence. Notably, MRD-S detection occurred before clinical or radiological detection of recurrence. These results highlight the potential of MRD to detect high-risk patients who could potentially benefit from earlier salvage local therapies, treatment intensification, or additional evaluation during the surveillance period.

Previously reported baseline MRD detection rates in patients with HPV- HNSCC range from 75% to 100% when using tumor-informed approaches in small cohorts^14,24,25^. This study represents the largest MRD study of patients with locally advanced HPV- HNSCC who were treated with surgery. Looking at this specific population helps assesses test characteristics in a specific use case to inform further investigations of how ctDNA can be incorporated into clinical care. MRD detection from surgery to the start of adjuvant treatment (MRD-E), was not associated with recurrence or survival in this cohort, likely due to the role of adjuvant treatment in preventing recurrence among patients with high-risk disease. Notably, of the eight patients in this cohort who underwent surgery alone and had an MRD-E result, seven developed recurrences. These findings strongly suggest that patients with positive post-operative MRD results could potentially benefit from intensification of adjuvant treatment, even if they do not have standard indications for adjuvant radiation and/or systemic therapy. Currently, multiple prospective trials are evaluating the potential of MRD-guided adjuvant therapy in solid tumors, including CRC, breast cancer, and locally advanced HNSCC^26–28^.

Post-treatment detection of MRD (MRD-TC) emerged as a strong predictor of disease relapse. This finding is consistent with prior work in other solid tumors, such as colorectal carcinoma (CRC), where the detection of ctDNA after adjuvant chemotherapy has been shown to predict recurrence^29–32^. The results of this study build on the emerging literature showing the utility of ctDNA-based MRD assays in LA-HNSCC. Two previously published small cohort studies evaluating ctDNA-based MRD assays in patients with LA-HNSCC showed comparable test characteristics to the results seen here^14,24^. Collectively, these results support further investigations to establish how ctDNA MRD assays can be used to guide patient care, as has been done in other tumor types, such as CRC, where ctDNA-guided adjuvant treatment was able to select patients for treatment deintensification without compromising RFS^29^.

As more data emerge supporting the prognostic value of MRD testing in HNSCC, the multifactorial effects on patient care should be considered and evaluated. For example, in endometrial cancer, another tumor type where surveillance requires invasive, uncomfortable physical exams, studies have demonstrated that some patients prefer MRD testing over physical examinations^33^. Research suggests that patients feel and believe MRD testing increases confidence when used alongside skin checks or imaging scans^33^, and a negative MRD result reduces anxiety about cancer recurrence in many patients with cancer. Some data suggest MRD assays may be able to reduce anxiety^34^, highlighting further potential benefits of ctDNA-based MRD assays in this population. Ongoing work to incorporate whole genome sequencing and methylation data to have the potential to improve test performances, which will further encourage use of MRD assays in clinical practice.

### Limitations

Our study has several limitations. As these specimens were collected during routine clinical care, the study did not employ power calculations or sample size justifications, and the lack of a standardized sample collection timeline led to a different sample size for the time windows analyzed. Additionally, patients were treated based on guidelines and multidisciplinary expertise rather than a prespecified protocol, which resulted in heterogeneous treatment plans (surgery with/without adjuvant therapy). The small sample size and low number of events in the analyzed groups resulted in imprecise effect size estimates and wide confidence intervals, which limit inference from these data. Additional limitations of this study include single site design and predominantly white, non-Hispanic male patient population. Despite these limitations, these findings support the continued study of tumor-informed, personalized MRD monitoring as part of curative-intent treatment in patients with locally-advanced HPV- HNSCC.

## Conclusion

These findings provide further support the prognostic values of ctDNA-based MRD assays among patients with LA-HNSCC. Future investigations should evaluate how MRD assays can be used in clinical care, particularly whether they can identify patients who may benefit from treatment intensification or deintensification and the role of these assays in post-treatment surveillance.

## Data Availability

All data produced in the present work are contained in the manuscript.

## Authors’ disclosures

G.B. F.Z. P.D. and S.J. were Predicine employees and owned Predicine stock during the study’s duration but did not influence the publication decision.

T.J.R. was a non-executive board director for Biocon Biologics Ltd and received research funding from Coherus Biosciences.

D.L.F receive research support from Calico, Predicine, BostonGene, NeoGenomics, and Haystack (Quest) and consulting fees from Merck, Noetic, Chrysalis Biomedical Advisors, NeoGenomics, Arcadia, Focus, GT Molecular and Olympus.

A.S.F. reports grant funding from Haystack Oncology, Inc.

D.A.R., J.M., S.N., R.D.M., V.E., D.T.L, J.J.P., M.A.V., D.G.D., A.L.F., J.D.R. report no disclosures.

## Authors’ Contributions

D.L.F: Conceptualization, investigation, methodology, writing–original draft, writing–review and editing. D.R.T: Data curation, software, formal analysis, methodology, writing–original draft, writing–review, and editing.

T.J.R: Conceptualization, investigation, methodology, investigation, writing–review and editing. G.B., F.Z., P.D., S.J.: Sample processing for PredicineBEACON MRD

J.M., S.N., V.E.: Plasma sample processing.

R.D.M, D.T.L, K.S.E., J.J.P, M.A.V., D.G.D., A.F., J.D.R.: Sample acquisition, writing-review and editing.

A.F.: Assessment of FFPE tumor sections, writing and editing.

## Funding Statement

This work was funded in kind by Predicine Laboratories and NIH/NIDCR R03DE030550 (Daniel L. Faden).

The sponsor did not play a role in the design and conduct study, data collection, data analysis, interpretation of the data; preparation, review, or approval of the manuscript; and decision to submit the manuscript for publication The sponsor did assist with data management and specimen analysis.

## Acknowledgments

This work was funded by in-kind assay support from Predicine Laboratories.

## Meeting Presentation

These results were submitted for presentation at the ASTRO/ASCO Multidisciplinary Head and Neck Symposium in Palm Desert, CA (Feb 19-21, 2026).

## Notes

### Competing Interest Statement

Associations with commercial entities that provided support for the work reported in the submitted manuscript (the timeframe for disclosure in this section of the form is the lifespan of the work being reported).

### Author Declarations

Dana-Farber/Harvard Cancer Center Institutional Review Board (DFCI 18-653) gave ethical approval for this work and all patients provided written informed consent.

