## Supplementary Figures 1-2 for "Prognostic value of tumor-informed ctDNA in HPV-independent head and neck squamous cell carcinoma"

Supplementary Figure 1: Baseline tumor fraction and analysis of tumor biopsies

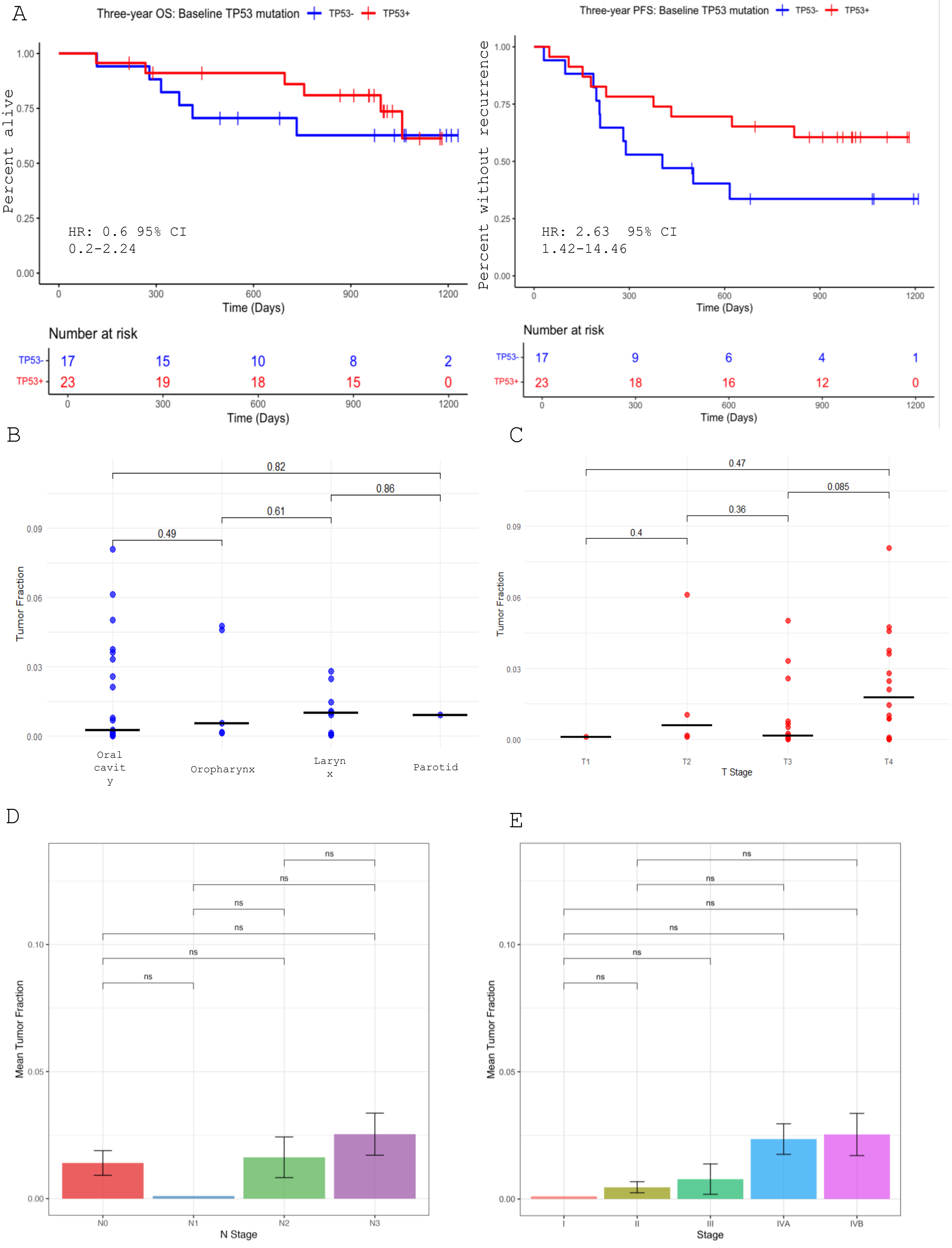

Supplementary Figure 1 (continued): Baseline tumor fraction and analysis of tumor biopsies

F

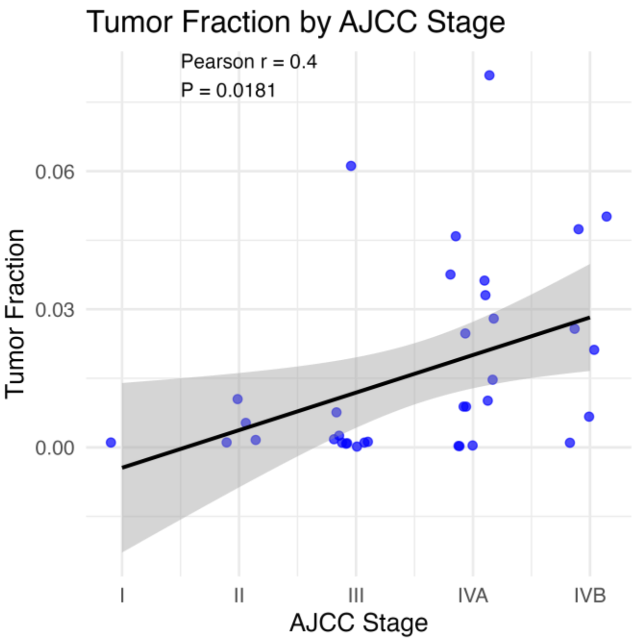


G

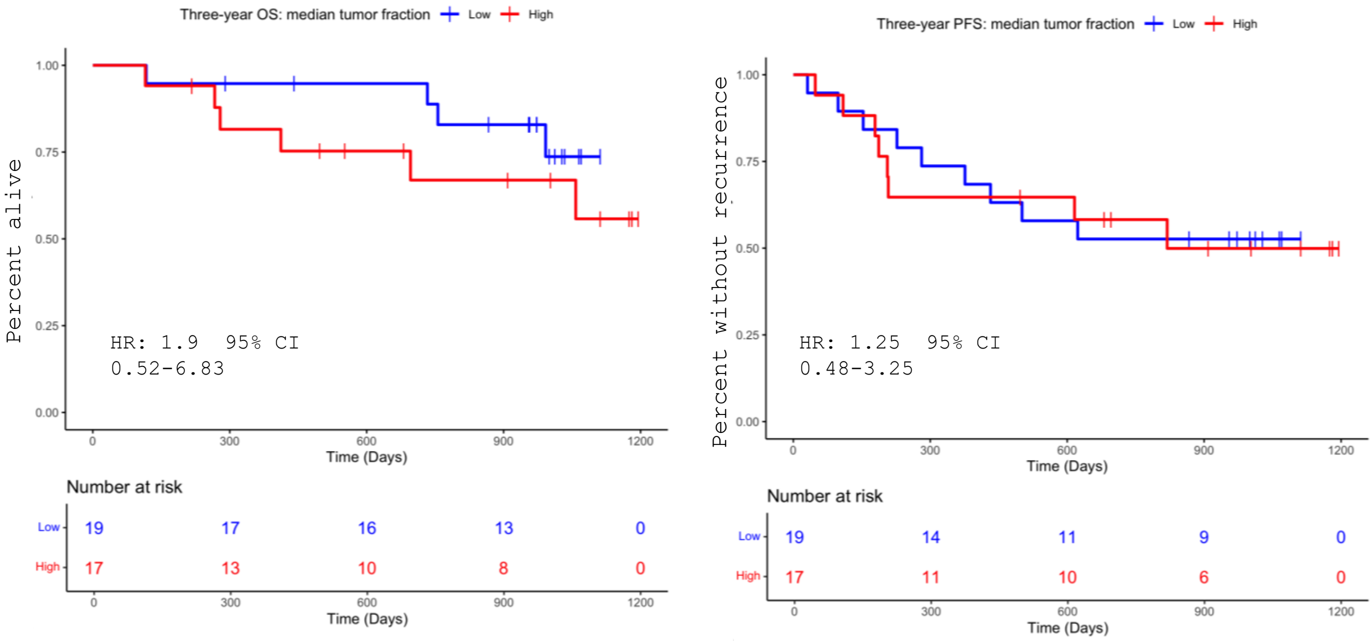

### Supplementary Figure 2A: OS and RFS by MRD-E status

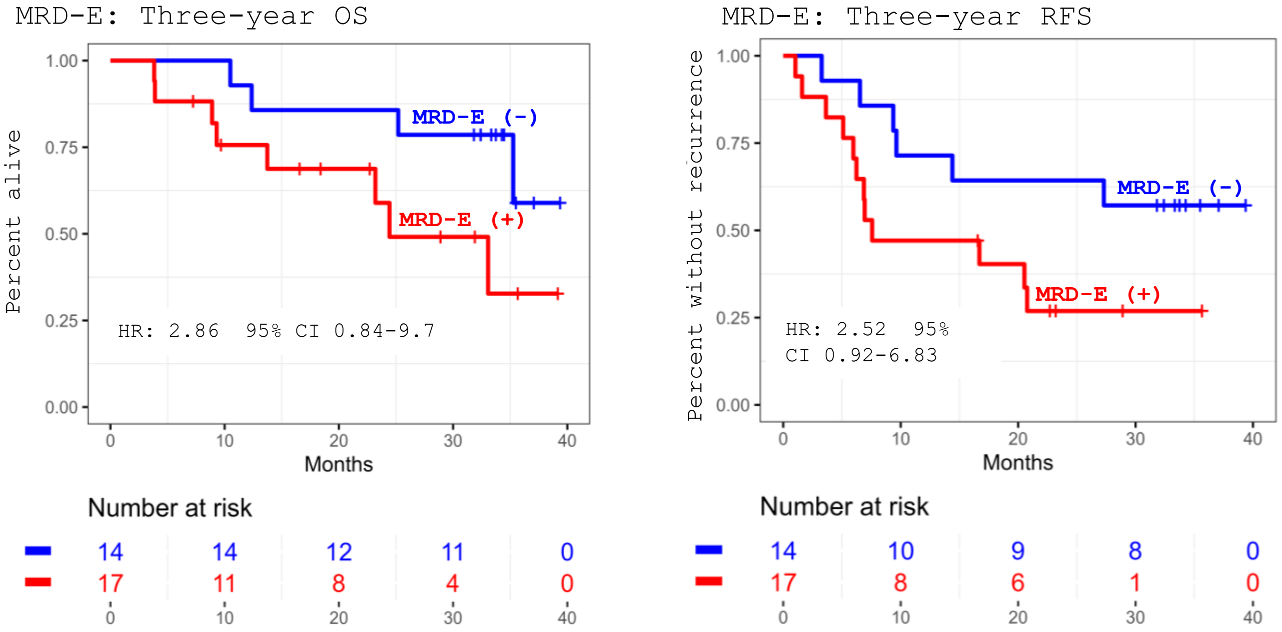


### Supplementary Figure 2B: OS and RFS for the entire cohort

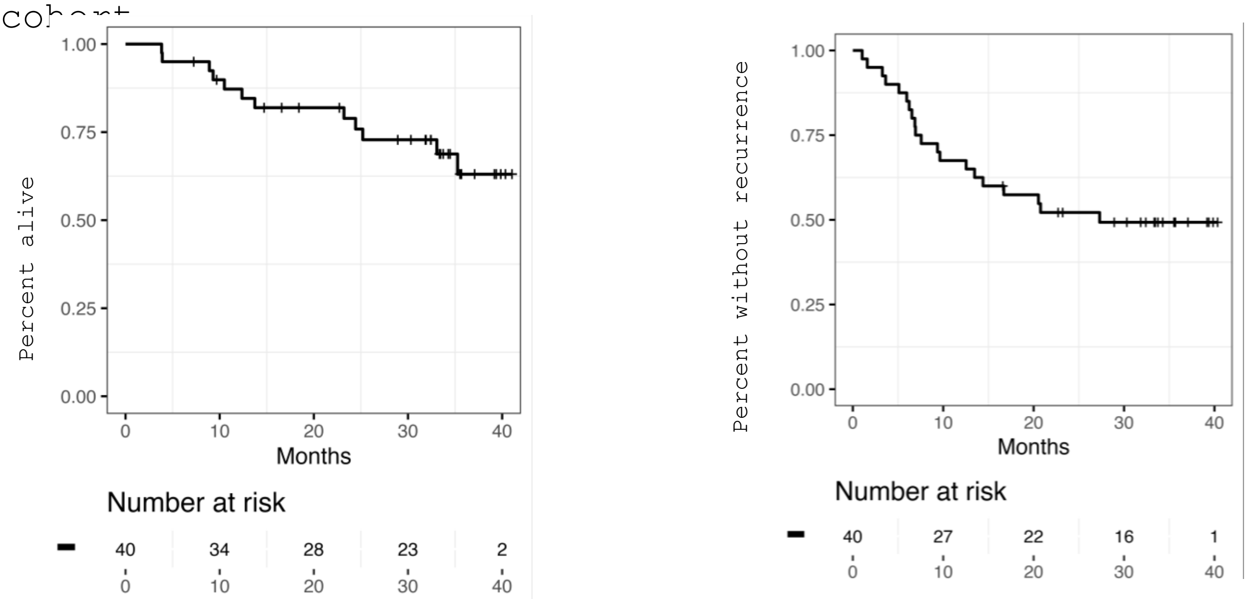


### Supplementary Figure 2 C. ENE

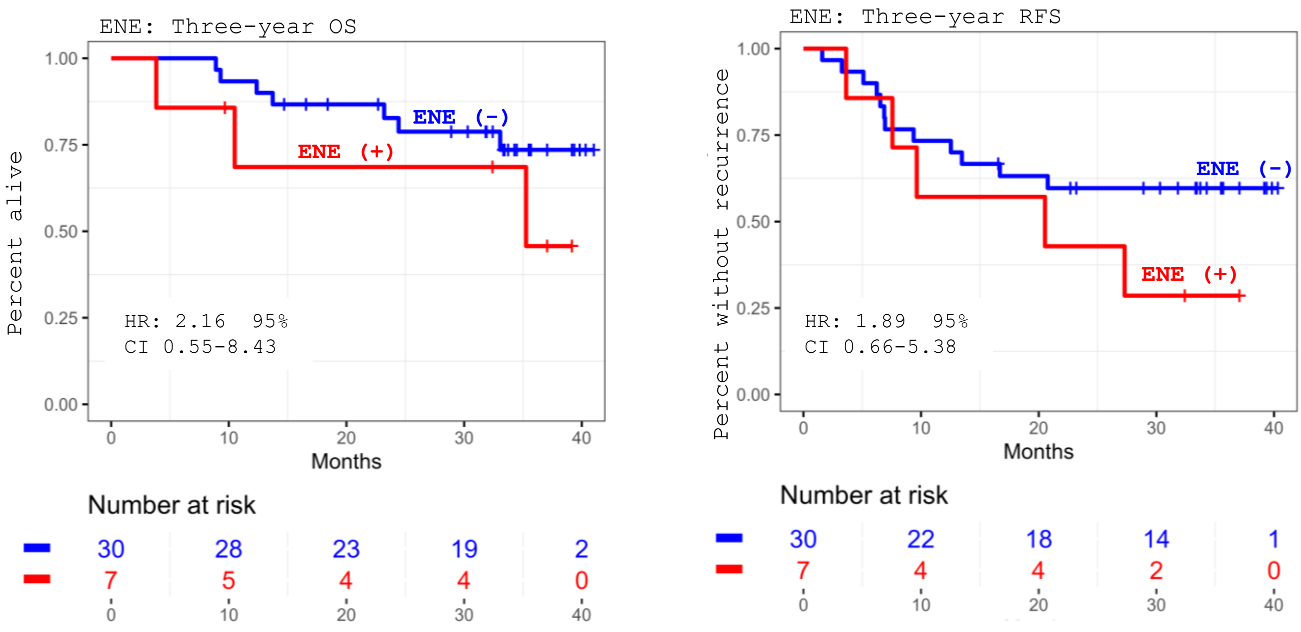

Supplementary Figure 2: RFS and OS by margin status and AJCC stage

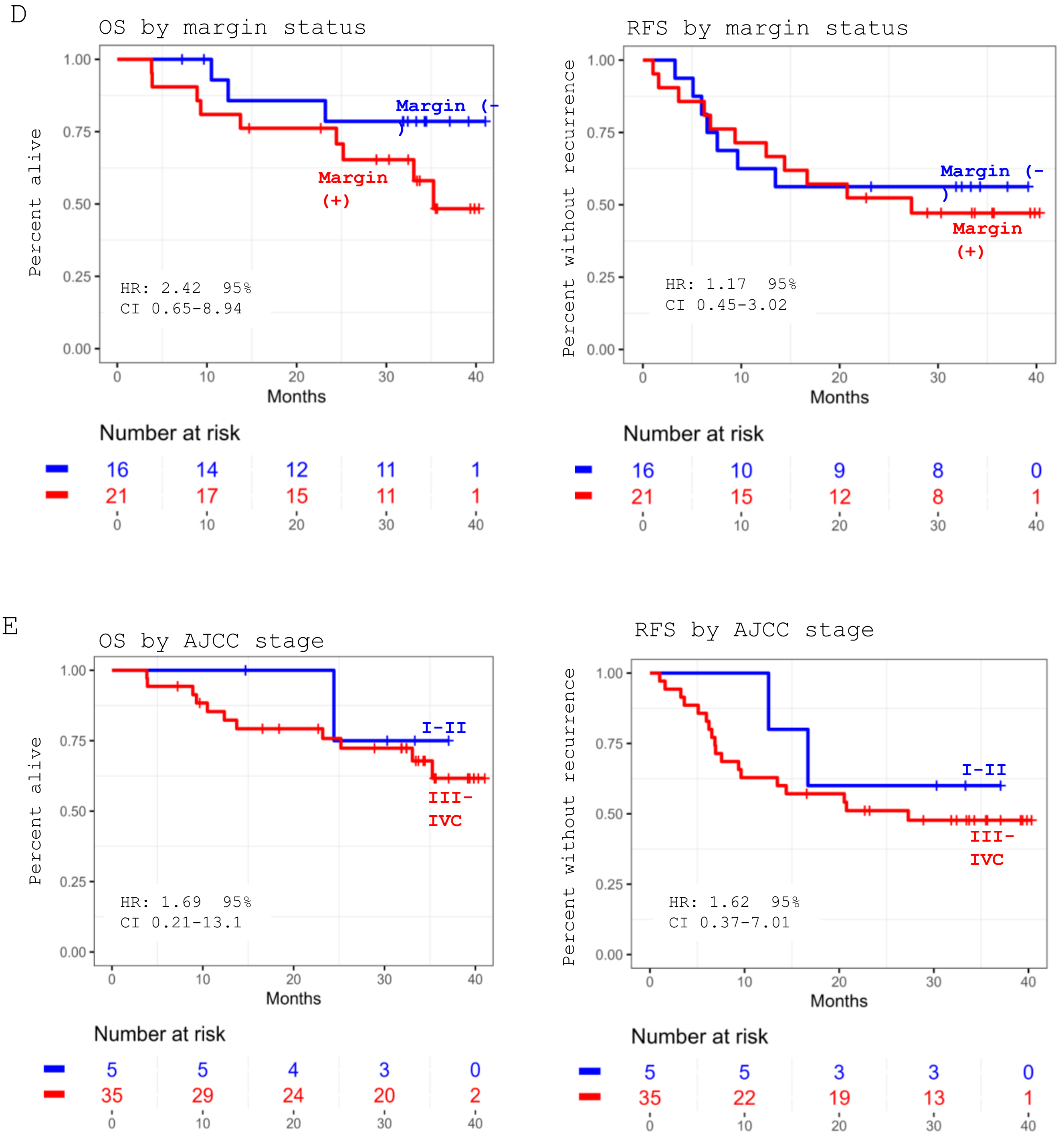
