## Supplementary Table 1 for "Prognostic value of tumor-informed ctDNA in HPV-independent head and neck squamous cell carcinoma"

**Supplementary Table and Figure Legends**

**Supplementary Table 1.** Sensitivity and specificity at a patient and test level during specific periods of follow-up. PPV: Positive predictive value, NPV: Negative predictive value.

| | | Patient level: Recurrence | | Test level: Recurrence | |
|---|---|---|---|---|---|
| | | **Yes** | **No** | **Yes** | **No** |
| **MRD-E** | **(+)** | 13 | 5 | 16 | 7 |
| | **(-)** | 4 | 10 | 13 | 21 |
| | **Sensitivity** | 77% | | 55% | |
| | **Specificity** | 67% | | 75% | |
| | **PPV** | 72% | | 70% | |
| | **NPV** | 71% | | 62% | |

| | | Patient level: Recurrence | | Test level: Recurrence | |
|---|---|---|---|---|---|
| | | **Yes** | **No** | **Yes** | **No** |
| **MRD-TC** | **(+)** | 7 | 1 | 7 | 1 |
| | **(-)** | 2 | 10 | 4 | 13 |
| | **Sensitivity** | 78% | | 64% | |
| | **Specificity** | 91% | | 93% | |
| | **PPV** | 88% | | 88% | |
| | **NPV** | 83% | | 77% | |

| | | Patient level: Recurrence | | Test level: Recurrence | |
|---|---|---|---|---|---|
| | | **Yes** | **No** | **Yes** | **No** |
| **MRD-S** | **(+)** | 8 | 2 | 10 | 2 |
| | **(-)** | 1 | 13 | 5 | 18 |
| | **Sensitivity** | 89% | | 67% | |
| | **Specificity** | 87% | | 90% | |
| | **PPV** | 80% | | 83% | |
| | **NPV** | 93% | | 78% | |

*MRD-E = minimal residual disease-early; MRD-TC = minimal residual disease – treatment completion; MRD-S = minimal residual disease during surveillance; PPV = positive predictive value; NPV = negative predictive value*

**Supplementary Figure 1**. Baseline tumor fraction and analysis of tumor biopsies. (A) Survival outcomes based on *TP53* mutation status. (B) Tumor fraction based on anatomical location. (C) Tumor fraction based on T stage, N stage (D), and 8$^{th}$ AJCC staging system (E). (F) Pearson correlation between stage and tumor fraction. (G) Survival outcomes based on median tumor fraction for the cohort.

**Supplementary Figure 2.** Survival outcomes based on different variables. (A) OS and RFS based on MRD-S status. (B) OS and RFS for all patients. (C) Survival outcomes based on the presence of extra-nodal extension (ENE), margins (D), or stage (I and II vs III and IV) (E).
